# Factors associated with deaths due to COVID-19 versus other causes: population-based cohort analysis of UK primary care data and linked national death registrations within the OpenSAFELY platform

**DOI:** 10.1101/2021.01.15.21249756

**Authors:** K Bhaskaran, SCJ Bacon, SJW Evans, CJ Bates, CT Rentsch, B MacKenna, L Tomlinson, AJ Walker, A Schultze, CE Morton, D Grint, A Mehrkar, RM Eggo, P Inglesby, IJ Douglas, HI McDonald, J Cockburn, EJ Williamson, D Evans, HJ Curtis, WJ Hulme, J Parry, F Hester, S Harper, D Spiegelhalter, L Smeeth, B Goldacre

## Abstract

**Background:** Mortality from COVID-19 shows a strong relationship with age and pre-existing medical conditions, as does mortality from other causes. However it is unclear how specific factors are differentially associated with COVID-19 mortality as compared to mortality from other causes.

**Methods:** Working on behalf of NHS England, we carried out a cohort study within the OpenSAFELY platform. Primary care data from England were linked to national death registrations. We included all adults (aged ≥18 years) in the database on 1^st^ February 2020 and with >1 year of continuous prior registration, the cut-off date for deaths was 9^th^ November 2020. Associations between individual-level characteristics and COVID-19 and non-COVID deaths were estimated by fitting age- and sex-adjusted logistic models for these two outcomes.

**Results:** 17,456,515 individuals were included. 17,063 died from COVID-19 and 134,316 from other causes. Most factors associated with COVID-19 death were similarly associated with non-COVID death, but the magnitudes of association differed. Older age was more strongly associated with COVID-19 death than non-COVID death (e.g. ORs 40.7 [95% CI 37.7-43.8] and 29.6 [28.9-30.3] respectively for ≥80 vs 50-59 years), as was male sex, deprivation, obesity, and some comorbidities. Smoking, history of cancer and chronic liver disease had stronger associations with non-COVID than COVID-19 death. All non-white ethnic groups had higher odds than white of COVID-19 death (OR for Black: 2.20 [1.96-2.47], South Asian: 2.33 [2.16-2.52]), but lower odds than white of non-COVID death (Black: 0.88 [0.83-0.94], South Asian: 0.78 [0.75-0.81]).

**Interpretation:** Similar associations of most individual-level factors with COVID-19 and non-COVID death suggest that COVID-19 largely multiplies existing risks faced by patients, with some notable exceptions. Identifying the unique factors contributing to the excess COVID-19 mortality risk among non-white groups is a priority to inform efforts to reduce deaths from COVID-19.

**Funding:** Wellcome, Royal Society, National Institute for Health Research, National Institute for Health Research Oxford Biomedical Research Centre, UK Medical Research Council, Health Data Research UK.

## Introduction

Severe acute respiratory syndrome coronavirus 2 (SARS-CoV-2) has infected tens of millions of people worldwide, causing substantial mortality.^1^ Numerous factors have emerged as being associated with a higher risk of severe outcomes and death from COVID-19.^2^ Mortality appears to rise exponentially with increasing age. Male sex, obesity, socioeconomic deprivation, and a number of comorbidities are also associated with higher risk.^3^ Substantial variation in mortality by ethnicity has also been observed in several studies.^4-6^ However, little evidence is available on how the factors associated with COVID-19 mortality compare with the factors associated with mortality from other causes, and hence the extent to which a person’s risk of dying from COVID-19 is likely to be governed by their broader mortality risk. We know that increasing age is the major risk factor for all-cause mortality. It is possible that COVID-19 simply multiplies everyone’s risk of death by a constant factor, or it could be that some factors have a different effect on COVID-19 deaths specifically.

A previous analysis of death registration data in England and Wales showed an exponential relationships between adult age and both rates of COVID-19 death (between March and June 2020), and pre-pandemic rates of all-cause mortality derived from life tables, with a slightly steeper age-mortality association for COVID-19 death.^7^ A study in preprint using UK Biobank data from before the current pandemic examined how demographic characteristics and non-communicable disease comorbidities were associated with deaths from infections versus other causes; the authors observed broadly similar patterns of risk for the two outcomes, though the magnitude of associations differed.^8^ However, it is unclear to what extent findings from pre-pandemic infection-related deaths can be used to draw conclusions about COVID-19.

To our knowledge, no study to date has directly compared factors associated with COVID-19 versus non-COVID deaths in the same cohort. We aimed to address this by conducting parallel analyses of COVID-19 and non-COVID death outcomes using population-based data from England within the OpenSAFELY platform.

## Methods

### Study design and study population

A retrospective cohort study was carried out within OpenSAFELY, a new data analytics platform in England created to address urgent COVID-19 related questions, which has been described previously.^3^ We used routinely-collected electronic data from primary care practices using The Phoenix Partnership (TPP) SystmOne software, covering approximately 40% of the population in England, linked to Office of National Statistics (ONS) death registrations. We included all adults (aged 18 years or over) alive and under follow-up on 1^st^ February 2020, and with at least one year of continuous GP registration prior to this date, to ensure that baseline data could be adequately captured. We excluded people with missing age, sex, or index of multiple deprivation, since these are likely to indicate poor data quality. For a secondary analysis of deaths prior to the pandemic, a second cohort was extracted comprising all adults alive and under follow-up on 1^st^ February 2019 and with at least one year of GP registration prior to that date (hereafter referred to as the “2019 cohort”).

### Outcome and covariates

The outcomes were COVID-19 death, and deaths from causes other than COVID-19 (hereafter “non-COVID death”). Cause of death was assigned using the underlying cause of death field (coded in ICD-10) in the death registration. COVID-19 death was defined as any death with the underlying cause coded as U07.1 (“COVID-19, virus identified”) or U07.2 (“COVID-19, virus not identified”).^9^ Non-COVID deaths comprised all other deaths; these were also further sub-divided into categories covering the most common causes of death, namely cancer (ICD-10 chapter C), cardiovascular disease (chapter I), respiratory (chapter J), dementia/Alzheimer’s disease (F00-03 or G30), and other (all other ICD-10 codes). Two sensitivity analyses were done to check that our findings were robust to the way COVID-19 deaths were defined: (i) only using the U07.1 (“virus identified”) code which would likely have higher specificity; (ii) counting a U07.1/U07.2 code anywhere on the death certificate as a COVID-19 death, in case of variation in how underlying causes were assigned. For the secondary analysis of deaths prior to the pandemic, the outcome was all-cause mortality; this was based on a record for death in primary care, because ONS death registration linkage for 2019 was not available.

Covariates considered in the analysis included age (grouped as 18-39, 40-49, 50-59, 60-69, 70-79 and ≥80 years for descriptive analysis), sex, ethnicity (White, Mixed, South Asian, Black, Other, categories from the UK census), obesity (categorised as class I [body mass index 30-34.9kg/m^2^], II [35-39.9kg/m^2^], III [≥40kg/m^2^]), smoking status (never, former, current), index of multiple deprivation quintile (derived from the patient’s postcode at lower super output area level), and comorbidities considered potential risk factors for severe COVID-19 outcomes. These were diagnosed hypertension, chronic respiratory diseases other than asthma, asthma (categorised as with or without recent use of oral steroids), chronic heart disease, diabetes (categorised according to the most recent glycated haemoglobin (HbA1c) recorded in the 15 months prior to 1^st^ February 2020), non-haematological and haematological cancer (both categorised by recency of diagnosis, <1, 1-4.9, ≥5 years), reduced kidney function (categorised by estimated glomerular filtration rate derived from the most recent serum creatinine measure (30-<60, 15-<30, <15 mL/min/1.73m^2^ or a record of dialysis), chronic liver disease, stroke, dementia, other neurological disease (motor neurone disease, myasthenia gravis, multiple sclerosis, Parkinson’s disease, cerebral palsy, quadriplegia or hemiplegia, and progressive cerebellar disease), organ transplant, asplenia (splenectomy or a spleen dysfunction, including sickle cell disease), rheumatoid arthritis/lupus/psoriasis, and other immunosuppressive conditions (permanent immunodeficiency ever diagnosed, or aplastic anaemia or temporary immunodeficiency recorded within the last year).

Information on all covariates was obtained by searching TPP SystmOne records prior to 1^st^ February 2020 (or prior to 1^st^ February 2019, for the 2019 secondary analysis cohort) for specific coded data, based on a subset of SNOMED-CT mapped to Read version 3 codes. All codelists, along with detailed information on their compilation are available at https://codelists.opensafely.org for inspection and re-use by the wider research community.

### Statistical analysis

Follow-up time for mortality in the main cohort was from 1^st^ February 2020 until 9^th^ November 2020, which was the last date for which mortality data were complete. Overall absolute risks of COVID-19 death and non-COVID death (by cause of death category) were calculated for each age group, and standardised by sex; this was done by fitting a multinomial logistic regression model with outcome levels of died (by cause of death category) versus alive at end of follow-up, including covariates of age group and sex, and predicting risks for each outcome under a 50:50 male:female split.

To estimate differential associations between potential risk factors and mortality from COVID or non-COVID causes, binary logistic regression models were then fitted, with outcomes of (i) COVID-19 death and (ii) non-COVID death (all non-COVID causes combined). For each outcome, separate age- and sex-adjusted models were fitted for each covariate, with age parametrised as a 4-knot restricted cubic spline in regression models. An additional model was fitted with age group and sex only, to provide odds ratios for age group (since model estimates for spline terms are difficult to interpret). For the secondary analysis using the 2019 cohort, a similar set of age- and sex-adjusted logistic models was fitted using a follow-up time period from 1^st^ February to 9^th^ November 2019, with the outcome of all-cause mortality, which by definition represents non-COVID mortality during this pre-pandemic time period.

In further secondary analyses, models for each outcome were also fitted, adjusting for all covariates simultaneously to identify independent associations between covariates and outcomes. In a post-hoc analysis driven by the striking pattern of results for ethnicity, further age- and sex-adjusted logistic models were fitted to explore ethnic differences in the odds of death from specific cause of death categories. Finally, to directly estimate the comparative association between individual-level factors and COVID-19 versus non-COVID deaths, we fitted age- and sex-adjusted logistic models only including people who died, with cause of death (COVID-19 versus non-COVID) as the binary outcome variable. In these analyses, odds ratios >1 indicate that a variable has a more positive association with COVID-19 death than with non-COVID death, and vice versa.

Multiple imputation (10 imputations) was used to account for missing ethnicity, with the imputation model including all covariates from the main model and an indicator for the outcome. Those with missing body mass index were assumed to be non-obese, and those with missing smoking data were assumed to be never-smokers; we did not use multiple imputation for these variables as they are expected to be missing not at random in UK primary care.^10^ In sensitivity analyses, we excluded individuals with missing data (complete case analysis).

### Information Governance and Ethics

NHS England is the data controller; TPP is the data processor; and the key researchers on OpenSAFELY are acting on behalf of NHS England. OpenSAFELY is hosted within the TPP environment which is accredited to the ISO 27001 information security standard and is NHS IG Toolkit compliant;^11,12^ patient data are pseudonymised for analysis and linkage using industry standard cryptographic hashing techniques; all pseudonymised datasets transmitted for linkage onto OpenSAFELY are encrypted; access to the platform is via a virtual private network (VPN) connection, restricted to a small group of researchers who hold contracts with NHS England and only access the platform to initiate database queries and statistical models. All database activity is logged. No patient-level data leave the platform; only aggregate statistical outputs leave the platform environment following best practice for anonymisation of results such as statistical disclosure control for low cell counts.^13^ The OpenSAFELY platform adheres to the data protection principles of the UK Data Protection Act 2018 and the EU General Data Protection Regulation (GDPR) 2016. In March 2020, the Secretary of State for Health and Social Care used powers under the UK Health Service (Control of Patient Information) Regulations 2002 (COPI) to require organisations to process confidential patient information for the purposes of protecting public health, providing healthcare services to the public and monitoring and managing the COVID-19 outbreak and incidents of exposure.^14^ Taken together, these provide the legal bases to link patient datasets on the OpenSAFELY platform. This study was approved by the Health Research Authority (REC reference 20/LO/0651) and by the LSHTM Ethics Board (ref 21863).

## Results

17,456,515 individuals were included in the analysis, of whom 17,063 died with COVID-19 listed as the underlying cause, while 134,316 died of other underlying causes (Figure 1). Demographic characteristics are shown in Table 1.

**Table 1:** Characteristics of the primary study cohort

|  |  | N (%) |
| --- | --- | --- |
| **N** |  | 17,456,515 (100.0) |
| **Age (yrs)** | 18-39 | 5,965,744 (34.2) |
|  | 40-49 | 2,877,525 (16.5) |
|  | 50-59 | 3,085,141 (17.7) |
|  | 60-69 | 2,421,249 (13.9) |
|  | 70-79 | 1,963,205 (11.2) |
|  | 80+ | 1,143,651 (6.6) |
| **Sex** | Male | 8,739,169 (50.1) |
|  | Female | 8,717,346 (49.9) |
| **Body mass index (kg/m<sup>2</sup>)\*** | Not obese | 13,621,506 (78.0) |
|  | 30-34.9 (Obese class I) | 2,419,268 (13.9) |
|  | 35-39.9 (Obese class II) | 940,080 (5.4) |
|  | ≥40 (Obese class III) | 475,661 (2.7) |
| **Smoking status\*** | Never | 8,739,756 (50.1) |
|  | Former | 5,747,053 (32.9) |
|  | Current | 2,969,706 (17.0) |
| **Ethnicity** | White | 11,163,018 (63.9) |
|  | Mixed | 172,141 (1.0) |
|  | South Asian | 1,027,068 (5.9) |
|  | Black | 343,094 (2.0) |
|  | Other | 323,893 (1.9) |
|  | Missing | 4,427,301 (25.4) |
| **Index of Multiple Deprivation** | 1 (least deprived) | 3,519,427 (20.2) |
|  | 2 | 3,555,666 (20.4) |
|  | 3 | 3,515,186 (20.1) |
|  | 4 | 3,491,534 (20.0) |
|  | 5 (most deprived) | 3,374,702 (19.3) |
| **Comorbidities** |  |  |
| Hypertension |  | 5,990,510 (34.3) |
| Chronic respiratory disease |  | 711,370 (4.1) |
| Asthma | With no oral steroid use | 2,484,264 (14.2) |
|  | With oral steroid use | 296,251 (1.7) |
| Chronic heart disease |  | 1,179,367 (6.8) |
| Diabetes | With HbA1c<58 mmol/mol | 1,053,215 (6.0) |
|  | With HbA1c≥58 mmol/mol | 491,874 (2.8) |
|  | With no recent HbA1c measure | 196,831 (1.1) |
| Cancer (non-haematological) | Diagnosed < 1 year ago | 81,070 (0.5) |
|  | Diagnosed 1-4.9 years ago | 237,331 (1.4) |
|  | Diagnosed ≥5 years ago | 547,778 (3.1) |
| Haematological malignancy | Diagnosed < 1 year ago | 8,878 (0.1) |
|  | Diagnosed 1-4.9 years ago | 28,130 (0.2) |
|  | Diagnosed ≥5 years ago | 64,022 (0.4) |
| Reduced kidney function | Estimated GFR 30-60 | 1,012,185 (5.8) |
|  | Estimated GFR 15-<30 | 60,836 (0.3) |
|  | Estimated GFR <15 or dialysis | 31,027 (0.2) |
| Chronic liver disease |  | 100,844 (0.6) |
| Dementia |  | 41,460 (0.2) |
| Stroke |  | 367,717 (2.1) |
| Other neurological disease |  | 172,055 (1.0) |
| Organ transplant |  | 20,194 (0.1) |
| Asplenia |  | 28,083 (0.2) |
| Rheumatoid arthritis/lupus/psoriasis |  | 885,573 (5.1) |
| Other immunosuppressive conditions |  | 45,307 (0.3) |

\*missing BMI included in 'not obese' (n = 3,711,186 (21.3%); missing smoking included in 'never smoker' (n = 732,342 (4.2%))

**Figure 1:**
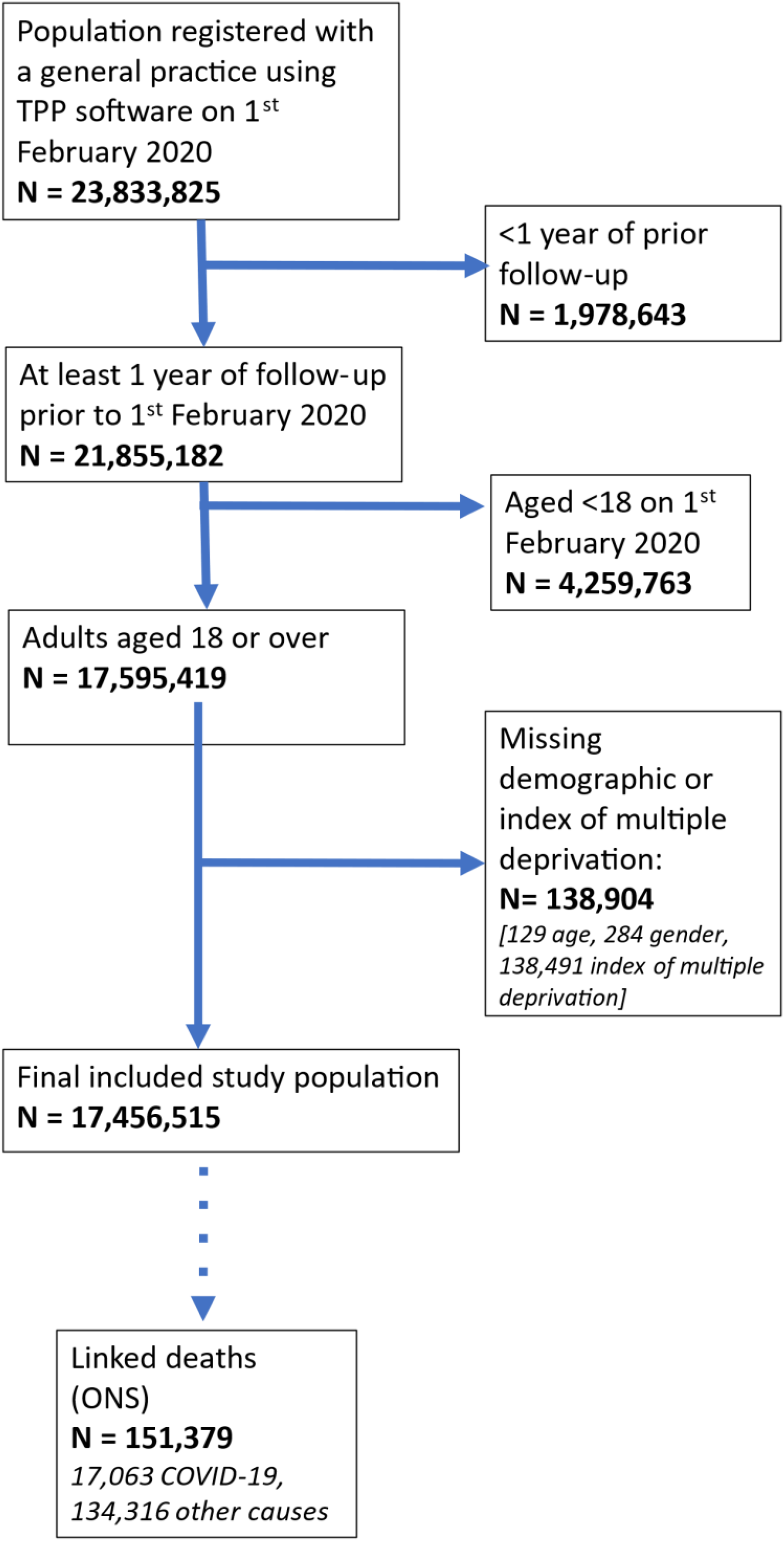
Flow chart of participants in the primary study cohort

As expected, the absolute risks of both COVID-19 death and death from other causes during the 283-day study period were highly dependent on age. The risks of COVID-19 death ranged from 0.00094% in those aged 18-39 years to 0.74% in those aged ≥80 years, and were similar to the risks of deaths from other respiratory causes combined (Figure 2, appendix Table A1). Risks of death from cancer and cardiovascular disease were higher (ranging from 0.0027% to 1.5% in the youngest to oldest age groups for cardiovascular disease, and from 0.0053% to 1.2% for cancer death); in those aged ≥80, the risk of dementia/ Alzheimer’s death was 1.5%.

**Figure 2:**
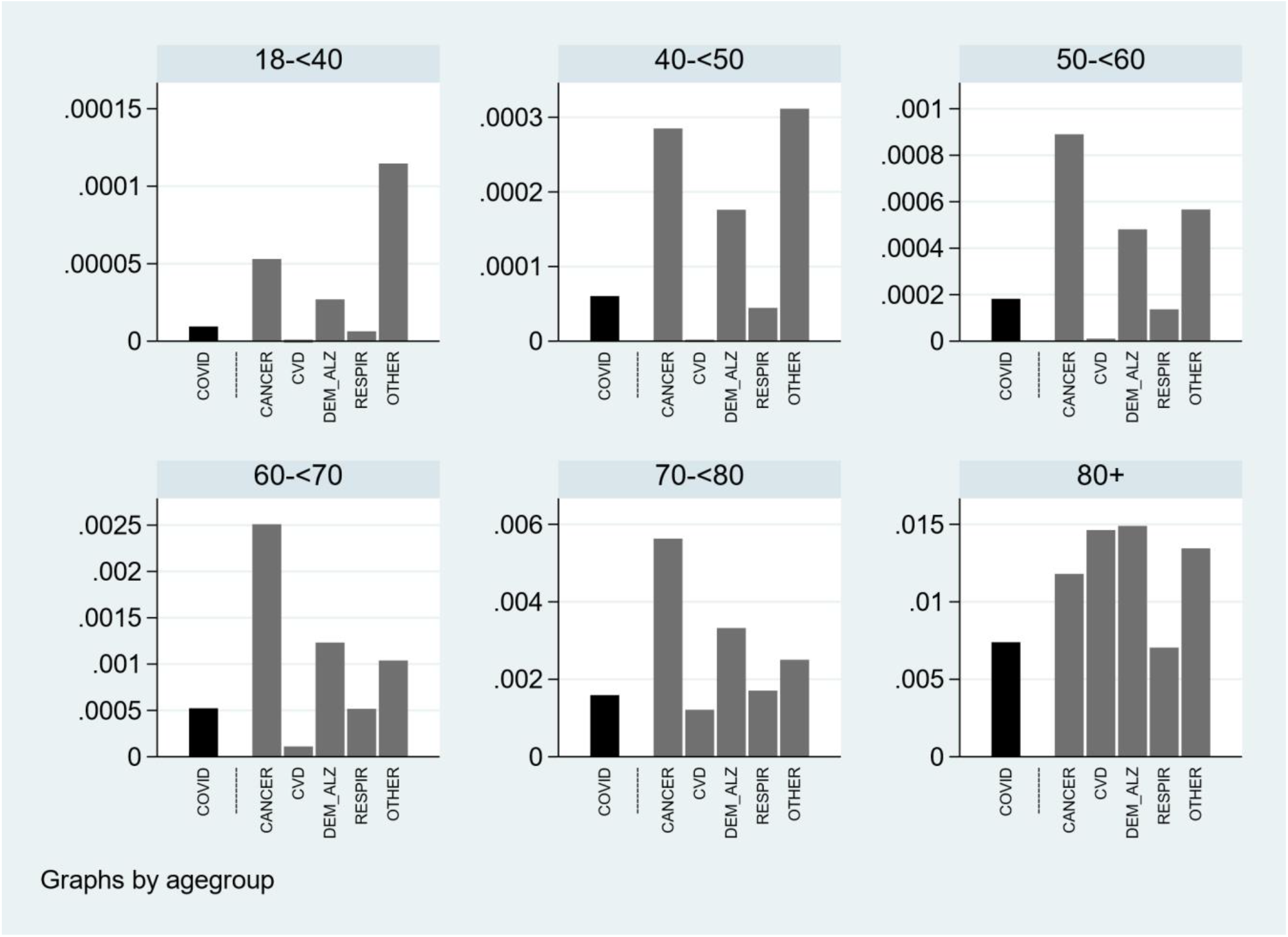
Absolute risk of death from different causes between 1^st^ February and 9^th^ November 2020, by age group <u>WARNING: please note the different y-scales in each panel</u> Absolute risks of death calculated estimated from a multinomial logistic regression model with alive versus died from specific causes as outcomes, and with age group and sex fitted as covariates; estimates are standardised to a 50% male/female gender balance within each age group.

Most individual-level factors associated with odds of COVID-19 death had qualitatively similar associations with non-COVID death, but the magnitudes of association differed (Figure 3). Age, male gender, obesity, deprivation, and some comorbidities (notably uncontrolled diabetes, severe asthma, dementia and organ transplant) had more pronounced positive associations with COVID-19 death than non-COVID death, while smoking, history of cancer and chronic liver disease had more pronounced positive associations with non-COVID death than COVID death.

**Figure 3:**
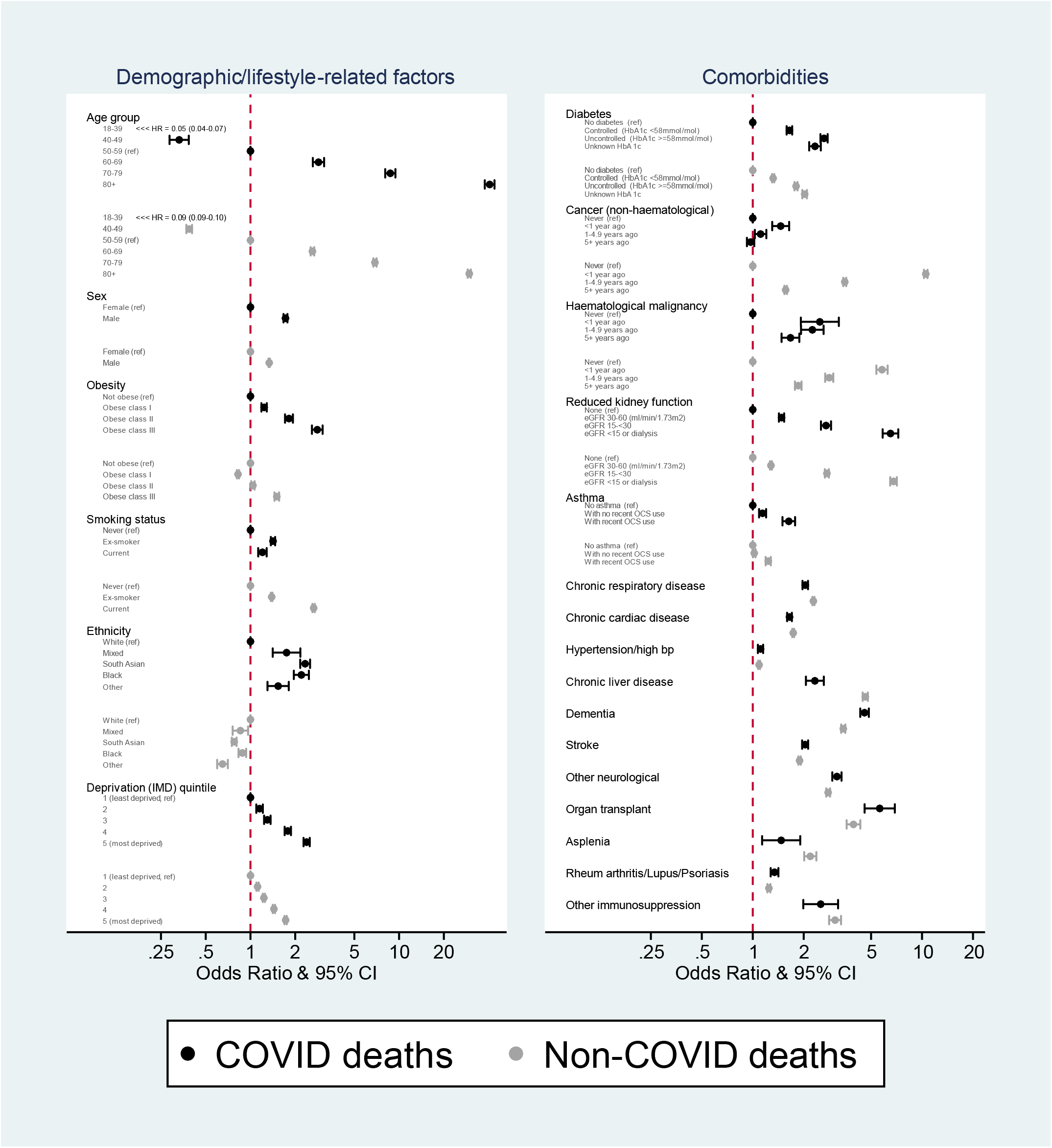
Odds ratios for the association between individual factors and COVID-19 and non-COVID mortality, adjusted for age and sex Estimates for each covariate were produced by fitting two age- and sex-adjusted logistic models with outcomes of COVID-19 death and death from other causes respectively

Non-white ethnicity had opposite associations with COVID-19 and non-COVID death: non-white ethnic groups had higher odds than white of COVID-19 death, but lower odds than white of death from non-COVID causes (Figure 3). This was also seen when non-COVID deaths were divided into more specific cause of death categories: non-white groups had similar or lower odds than white of death from cancer, cardiovascular disease, dementia/Alzheimer’s, and respiratory causes of death (Figure 4).

**Figure 4:**
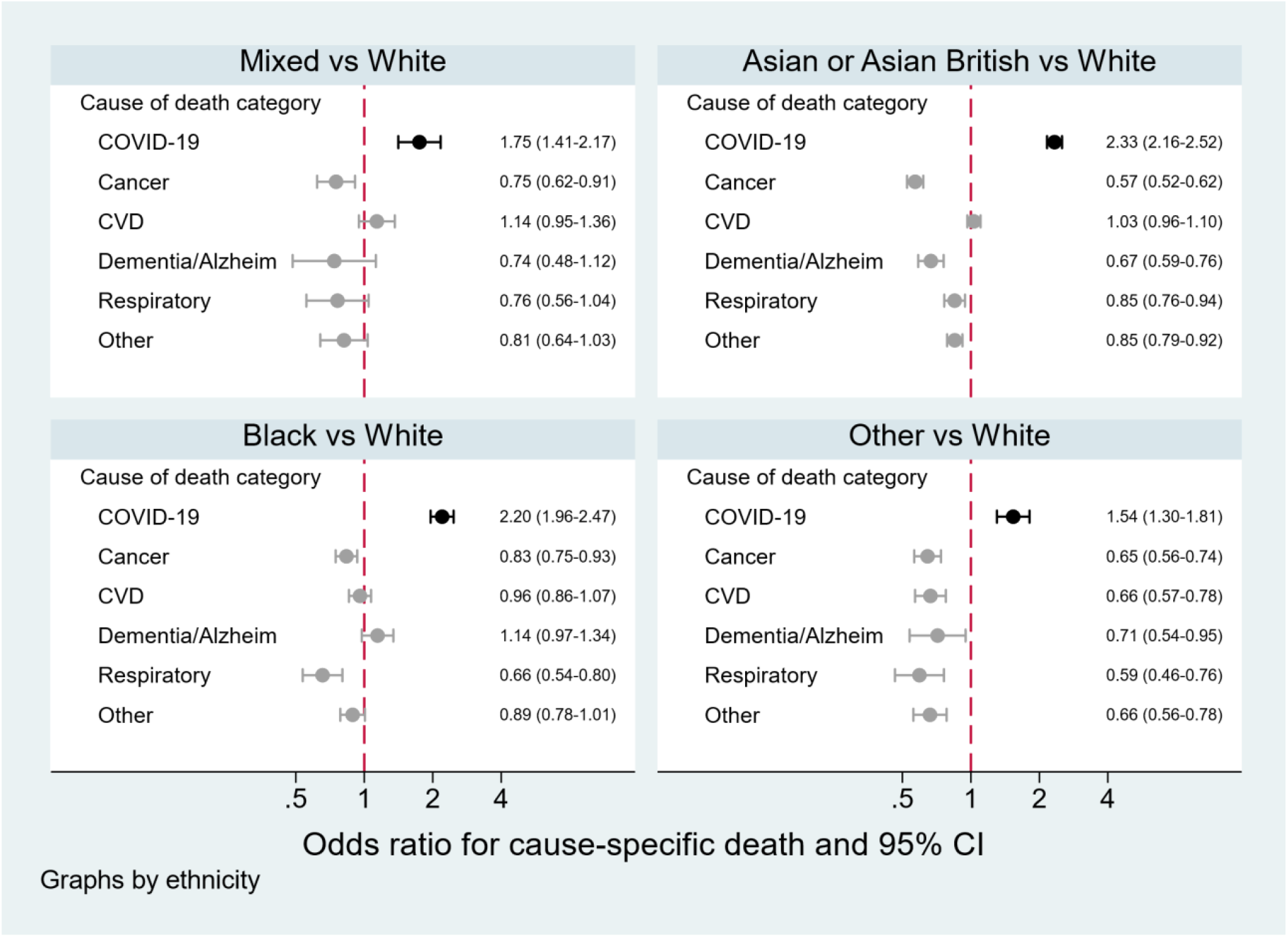
Odds ratios for the association between ethnicity and COVID-19 death and death from specific other causes, adjusted for age and sex From separate logistic regression models for each cause-specific mortality outcome, with age (spline), sex and ethnicity as covariates. Note: the dementia/Alzheimer’s outcome model was restricted to those aged ≥40y due to non-convergence when younger people were included

An analysis restricted to those who died, directly modelling the odds of COVID-19 versus non-COVID cause of death, confirmed the pattern of results from our primary analyses (Figure 5). Older age, male sex, obesity, ethnicity, higher deprivation, diabetes, reduced kidney function, asthma, stroke, dementia, other neurological conditions, history of organ transplant, and autoimmune diseases were to varying degrees stronger risk factors for COVID-19 deaths, while smoking, haematological and non-haematological cancers, chronic liver disease and asplenia were stronger risk factors for non-COVID death.

**Figure 5:**
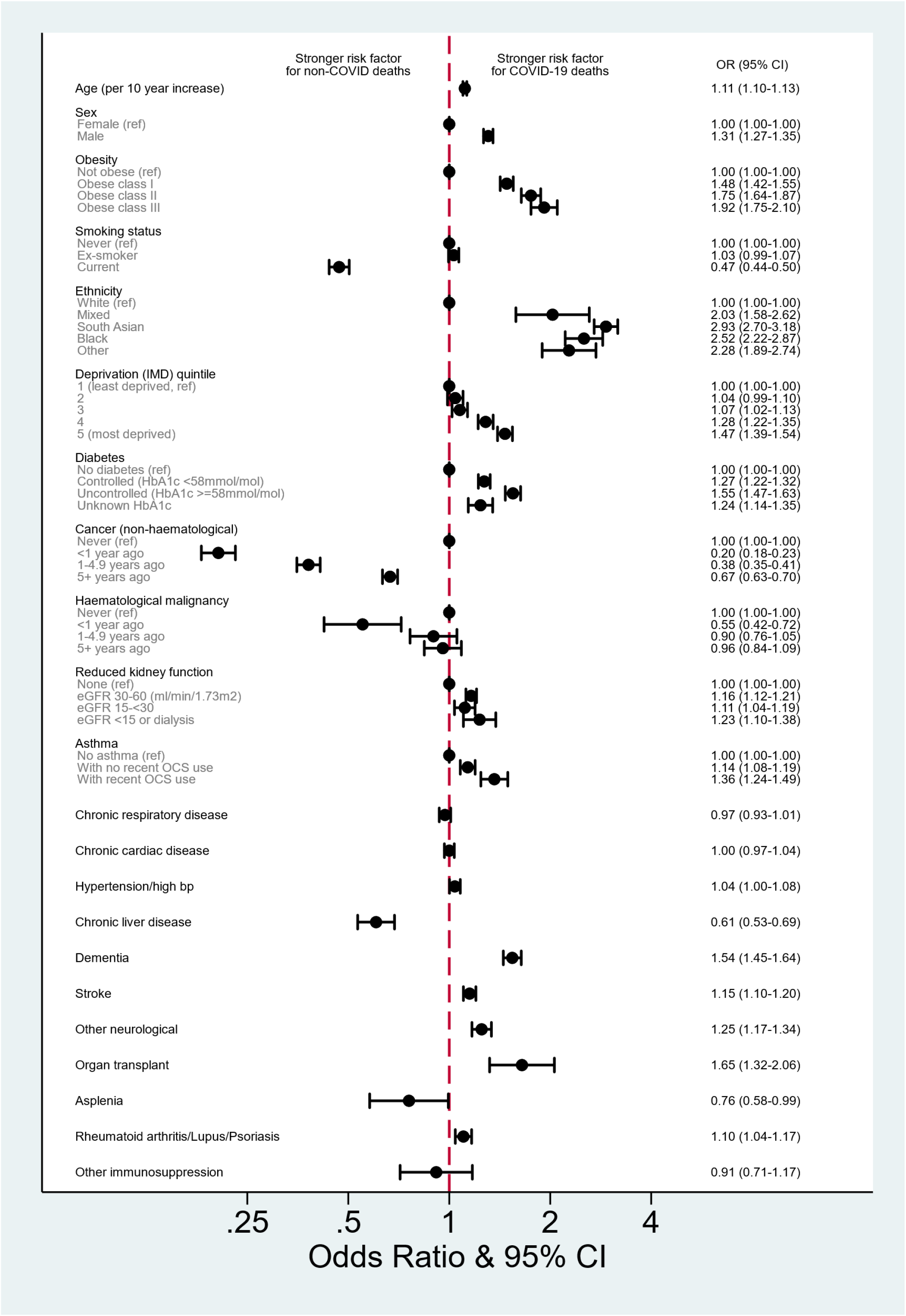
Odds ratio for COVID-19 cause of death (versus non-COVID causes) among those who died Estimates are from individual age and sex-adjusted logistic regression models for each factor of interest, including only individuals that died, and with an outcome of COVID-19 cause of death. Age was parameterised as a 4-knot restricted cubic spline in all models, except to estimate the effect of age itself, where a linear age term was used for ease of interpretation.

Associations between individual-level factors and non-COVID deaths were similar when deaths from 2019 were used to represent non-COVID deaths, and patterns of results were also similar in analyses mutually adjusted for all variables (appendix Table A2). In sensitivity analyses, varying how COVID-19 deaths were defined and using complete case analysis to deal with missing data made little difference to the results (appendix Figure A1a-e).

## Discussion

### Key findings

Patterns of association between individual-level factors and risk of COVID-19 death largely mirrored those for non-COVID deaths, suggesting that COVID-19 has largely acted to multiplying existing mortality risks faced by patients. However, there were notable exceptions. People from non-white ethnic groups were at substantially raised risk of COVID-19 death compared with white people, despite having similar or lower risks of deaths from other causes. Several other demographic characteristics, lifestyle-related factors and comorbidities had qualitatively similar associations with risk of both COVID-19 and non-COVID death but with different magnitudes of association: age, male sex, obesity, deprivation and some comorbidities including severe asthma, uncontrolled diabetes, dementia and organ transplant, had stronger associations with COVID-19 deaths than non-COVID deaths. The opposite was true of smoking and other comorbidities including cancers and chronic liver disease, which were more strongly associated with non-COVID deaths. During the period from February to November 2020, COVID-19 was a common cause of death in England, though the incidence of cancer and cardiovascular disease deaths was higher in all age groups, and the incidence of deaths recorded as being due to dementia/Alzheimer’s disease was also higher in the oldest individuals.

### Findings in the context of other evidence

Although individual level factors were generally similarly associated with COVID-19 and non-COVID death, some of the observed differences were striking, including the discrepant effects of ethnicity on the two outcomes. A lower overall mortality risk in Black and minority ethnic (BAME) groups has been observed before in an analysis of linked death registration data from 2001-2013 in Scotland, with suggested reasons including self-selection of healthy individuals among migrants, and healthier lifestyles and behaviours among BAME groups.^15^ A study in preprint using pre-pandemic data from UK Biobank also observed a reduced risk of mortality in BAME groups that was consistent for both infectious and non-infectious deaths;^8^ UK Biobank participants are not representative of the broader UK population, with evidence of a healthy volunteer selection bias,^16^ and it is possible that this bias may have operated more strongly in non-white ethnic groups. Nevertheless the evidence from both the internal comparison in the present study, and related data from other studies, suggests that the higher risk of poor COVID-19 outcomes reflects unique features of the pandemic rather than a generalised higher risk of death in non-white groups. Reasons might include a high likelihood of working in at-risk occupations with high exposure risk, such as health and social care, hospitality and public transportation; and a high likelihood of living in large, high-density or multigenerational households, which might act individually or in combination to increase the risk of infection, and thus the overall risk of COVID-19 death, particularly if a high exposure risk in younger people leads to increased infection in older people via households and community settings.^17^

Another notable finding was that the positive association between current smoking and COVID-19 death was substantially smaller than for non-COVID death. Current smokers also had a slightly smaller risk of COVID-19 death than ex-smokers, but the association was as expected for non-COVID death (current smokers at substantially higher risk than ex-smokers) suggesting that the finding was not driven by exposure misclassification or model mis-specification. Other studies have found smoking to be a significant risk factor for mortality among those hospitalised for COVID-19.^18^ If smokers have a raised risk of severe disease and death once infected, then it is possible that this has been diluted by a lower risk of infection in our general population-based study, but at present there is limited evidence to assess possible mechanisms that might explain such a reduced infection risk among smokers. Of note, the UK Biobank study found smoking to be more strongly associated with infection deaths than non-infection deaths in a pre-pandemic time period.^8^

Among comorbidities, history of non-haematological cancer stood out as having a substantially smaller association with COVID-19 death compared with non-COVID death; cancer patients and survivors are likely to have a high long-term risk of cancer recurrence driving a raised risk of non-COVID (cancer) death; our results suggest that this is proportionately larger than the more modest raised risk of COVID-19 death that might arise from compromised immunity or risk of infection complications. Any underlying raised COVID-19 risk in this group may have been mitigated if cancer patients and survivors were more likely to shield and/or be compliant with social distancing measures. The difference between COVID-19 and non-COVID mortality risk was less stark in haematological cancer survivors, in keeping with evidence that these cancers are likely associated with a larger raised risk of COVID-19 death.^3,19^

### Strengths and limitations

Study strengths include the large size of the study, providing high statistical power to investigate associations between a wide range of factors and mortality. Our use of routinely collected primary care data meant that information on a wide range of longitudinal, detailed patient characteristics and comorbidities were available, with individual-level linkage to death registrations providing near-complete ascertainment of mortality. Our findings were robust in a number of sensitivity analyses.

There were also some limitations. Comorbidity ascertainment relied on conditions being coded in the primary care record; conditions will only be coded when patients consult, which may not happen for early-stage or mild illness. Conversely, acute conditions requiring hospitalisation may have been missed if feedback from hospital to primary care providers was imperfect. Missing data was an issue for some variables, notably ethnicity and body mass index. We used multiple imputation to deal with missing ethnicity, and our findings were robust to an alternative “complete case analysis” approach. Recording of body mass index in primary care is highly likely to be missing not at random, violating a key assumption required for multiple imputation,^10^ so we instead assumed those with missing data to be non-obese, since obese individuals are more likely to have their weight recorded; however this could have caused some misclassification. Our results were again robust to an alternative “complete case approach”. We used the underlying cause of death field from the death registration to assign deaths as being due to COVID-19 or other causes, though results were similar in a sensitivity analysis where a COVID-19 death was defined based on a COVID-19 code anywhere on the death certificate. There may have been some misclassification of cause of death. Early in the epidemic, some COVID-19 deaths may have been misclassified as being due to other causes; for example a proportion of deaths due to COVID-19 occurring in care homes may have been misclassified as being due to dementia/Alzheimer’s, biasing downwards our estimate of the absolute contribution of COVID-19 to overall mortality.^20^ Conversely, during peak epidemic periods, a bias in the opposite direction may have occurred, if COVID-19 were entered as the presumed cause of death in uncertain cases. However, 16,049/17,063 COVID-19 deaths (94%) had the ICD-10 code U07.1 (“COVID-19 – virus identified”) implying that infection was confirmed by laboratory testing, so any misclassification in this direction is likely to have been minimal, and results were unchanged in a sensitivity analysis using an outcome definition restricted to this code. We were further reassured given that we found patterns of results for non-COVID deaths in 2020 to be similar to those for pre-pandemic deaths in the equivalent period of 2019. COVID-19 mortality may have been affected by the competing risk of non-COVID death, and vice versa; our use of logistic regression over a fixed time period effectively accounts for competing risks in a similar way to Fine & Gray modelling in a time to event framework,^21^ with our odds ratios incorporating any associations between covariates and outcome that are driven by the competing outcome.

### Implications for public health and research

In all age groups, absolute risk of COVID-19 death was approximately similar to the risk of death from other respiratory causes combined, but lower than the risk of death from cancer, cardiovascular disease and (among older individuals) dementia/Alzheimer’s. This highlights the importance of maintaining care and prevention services targeting these high-burden non-COVID conditions. Nevertheless, it should be remembered that the absolute risks of COVID-19 deaths we observed were in the context of major national efforts to suppress infection rates and targeted shielding for particular risk groups. The relative contribution of COVID-19 to mortality would undoubtedly have been higher in the absence of these policies. The broadly similar relationships between most individual-level factors and COVID-19 and non-COVID mortality suggest that COVID-19 largely amplifies a person’s underlying mortality risk (by a factor that will depend on current levels of circulating virus) based on their characteristics and medical history; public health decisions requiring prioritisation of vulnerable subgroups can therefore be informed by our knowledge of pre-pandemic mortality risks based on established risk factors. However there were important exceptions to this overall pattern. Understanding the drivers of uniquely raised risks of COVID-19 death associated with non-white ethnicity is a clear research priority; research into the role of occupations involving high levels of public contact, population density, and household composition will be key in further exploring the reasons, and thus informing mitigation strategies. Cancers were notably more strongly associated with non-COVID than COVID-19 deaths; a similar pattern was seen for chronic liver disease. However, this finding should not affect policies aimed at reducing risk in these groups, as even given a high underlying risk of non-COVID death, COVID-19 represents a potentially avoidable addition to the mortality risk. Emerging risk prediction tools can help to quantify this excess risk for individuals with different combinations of risk factors, and thus inform the targeting of mitigation measures, including prioritisation for forthcoming vaccines.^22,23^

### Conclusion

Demographic characteristics, lifestyle-related factors and comorbidities generally had qualitatively similar relationships with the risks of both COVID-19 death and non-COVID deaths, with some differences in the magnitudes of association. People from non-white ethnic groups had higher risks of COVID-19 death than white people, contrasting with similar or lower risks of deaths from other causes. This strongly suggests there are risk factors for mortality, specific to COVID-19, that are disproportionately affecting non-white groups; various factors along the causal chain culminating in COVID-19 death might contribute to the raised risks in non-white groups, including risk of exposure to the virus, risk of infection once exposed, underlying health and susceptibility to severe disease, health-seeking behaviour and health care received. Improved understanding of these factors is needed in order to tackle the increased mortality from COVID-19 among non-white groups. In conclusion, COVID-19 appears to largely act as if multiplying existing risks faced by patients, with the notable exceptions described in this paper.

## Data Availability

All data were linked, stored and analysed securely within the OpenSAFELY platform https://opensafely.org/. Data include pseudonymized data such as coded diagnoses, medications and physiological parameters. No free text data are included. All code is shared openly for review and re-use under MIT open license (https://github.com/opensafely/covid-vs-noncovid-deaths-research). Detailed pseudonymised patient data is potentially re-identifiable and therefore not shared. We rapidly delivered the OpenSAFELY data analysis platform without prior funding to deliver timely analyses on urgent research questions in the context of the global Covid-19 health emergency: now that the platform is established we are developing a formal process for external users to request access in collaboration with NHS England; details of this process will be published shortly on OpenSAFELY.org.

## Author contributions

BG conceived the OpenSAFELY platform and the approach. BG and LS lead the project overall and are guarantors. KB and SJWE led on study design. KB did the analysis and wrote the first draft. SB led on software development. AM led on information governance. Other contributions were – data curation - CB, JP, JC, SH, SB, DE, PI and CEM; disease category conceptualisation and codelists - KB CTR BM CB JC CEM AJW HIM IJD HJC JP; statistical analysis code: KB, EW; ethical approvals - HJC EW LS BG; software - SB, DE, PI, AJW, WJH, CEM, CB, FH, JC; writing (reviewing and editing) - KB, CTR, LT, BM, AS, AM, RME, CEM, IJD, SJWE, DS, LS, BG. All authors were involved in design and conceptual development and reviewed and approved the final manuscript.

## Patient and Public Involvement

Patients were not formally involved in developing this specific study design that was developed rapidly in the context of a global health emergency. We have developed a publicly available website https://opensafely.org/ through which we invite any patient or member of the public to contact us regarding this study or the broader OpenSAFELY project.

## APPENDIX

**Table A1:**
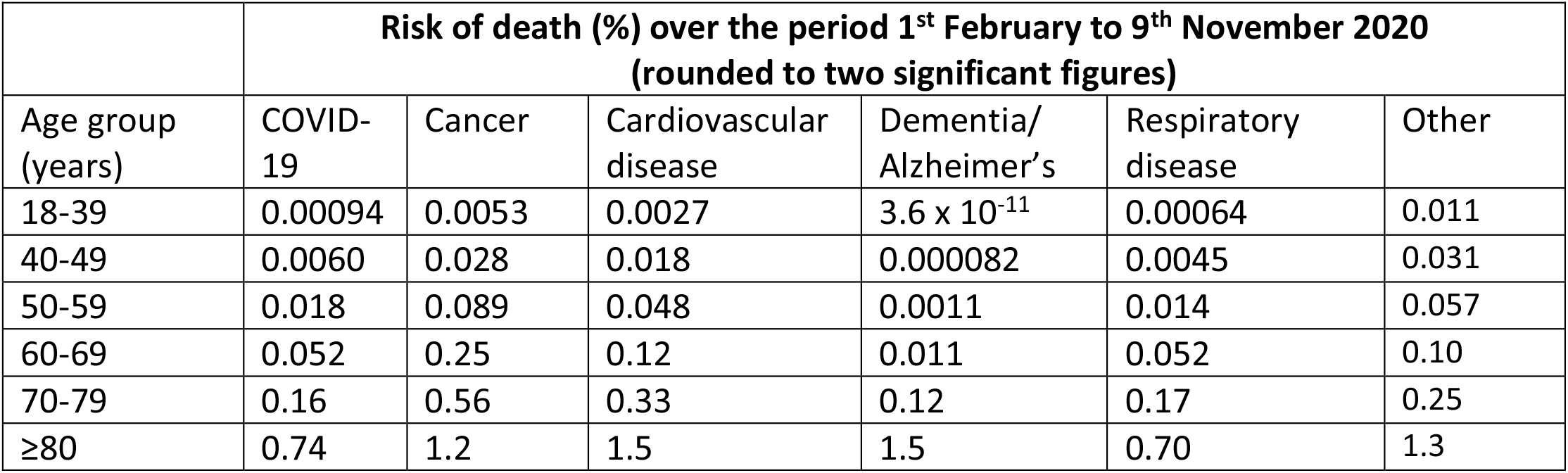
Estimated absolute risks of death from COVID-19 and non-COVID causes, stratified by age group

**Table A2:**
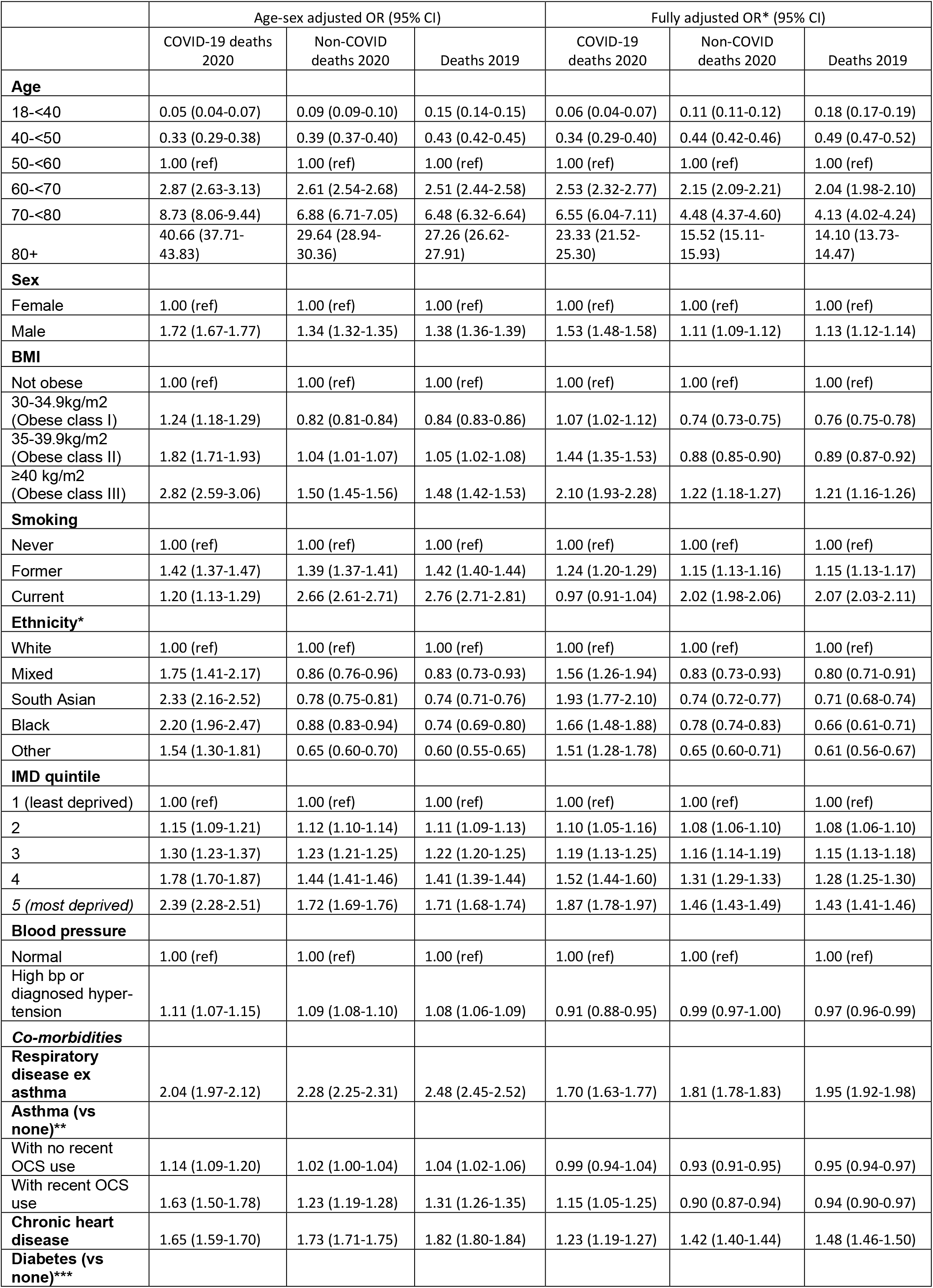
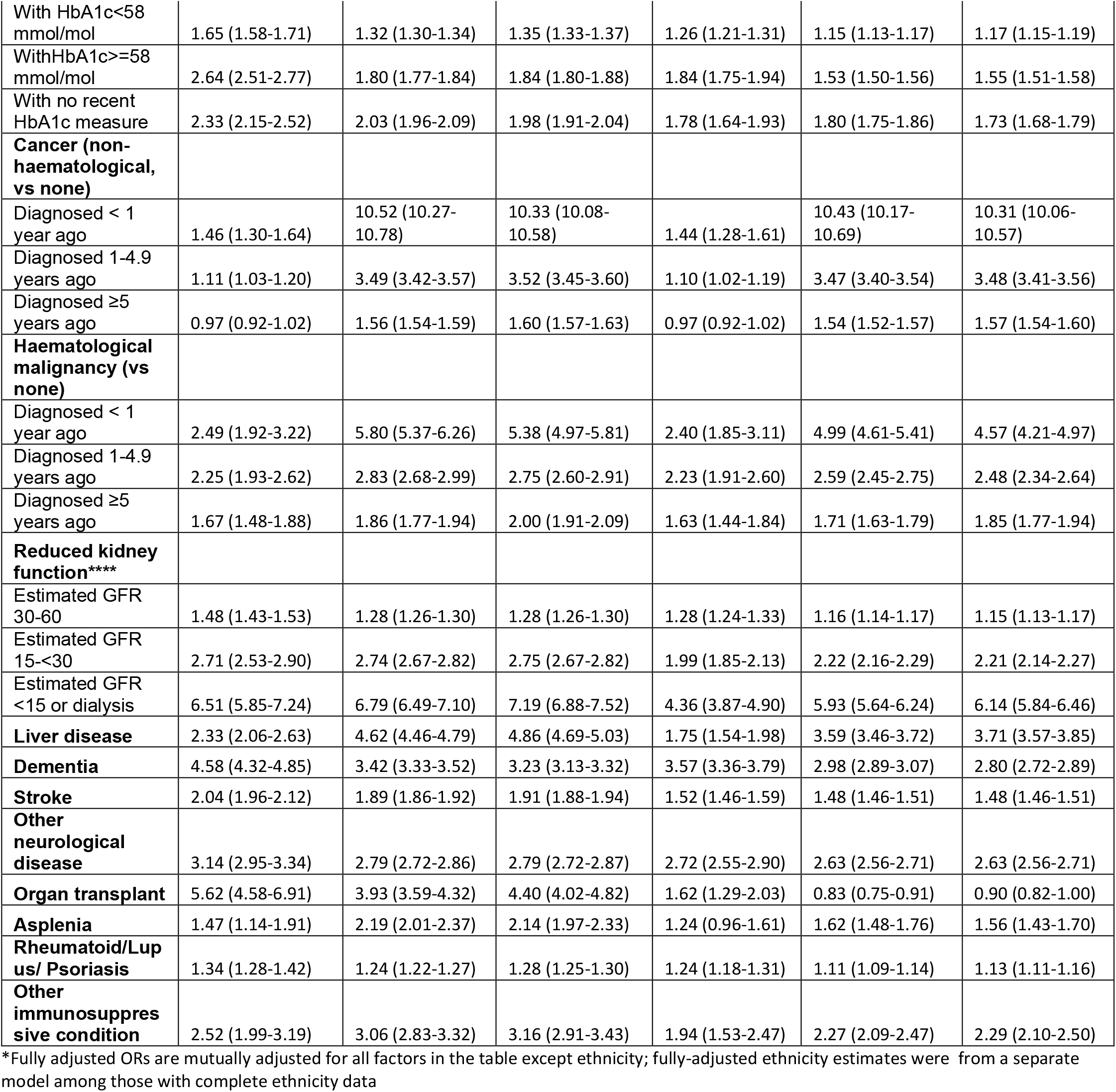
Age-sex adjusted and fully adjusted odds ratios for associations between individual-level factors and COVID-19/non-COVID deaths

**Figure A1:**
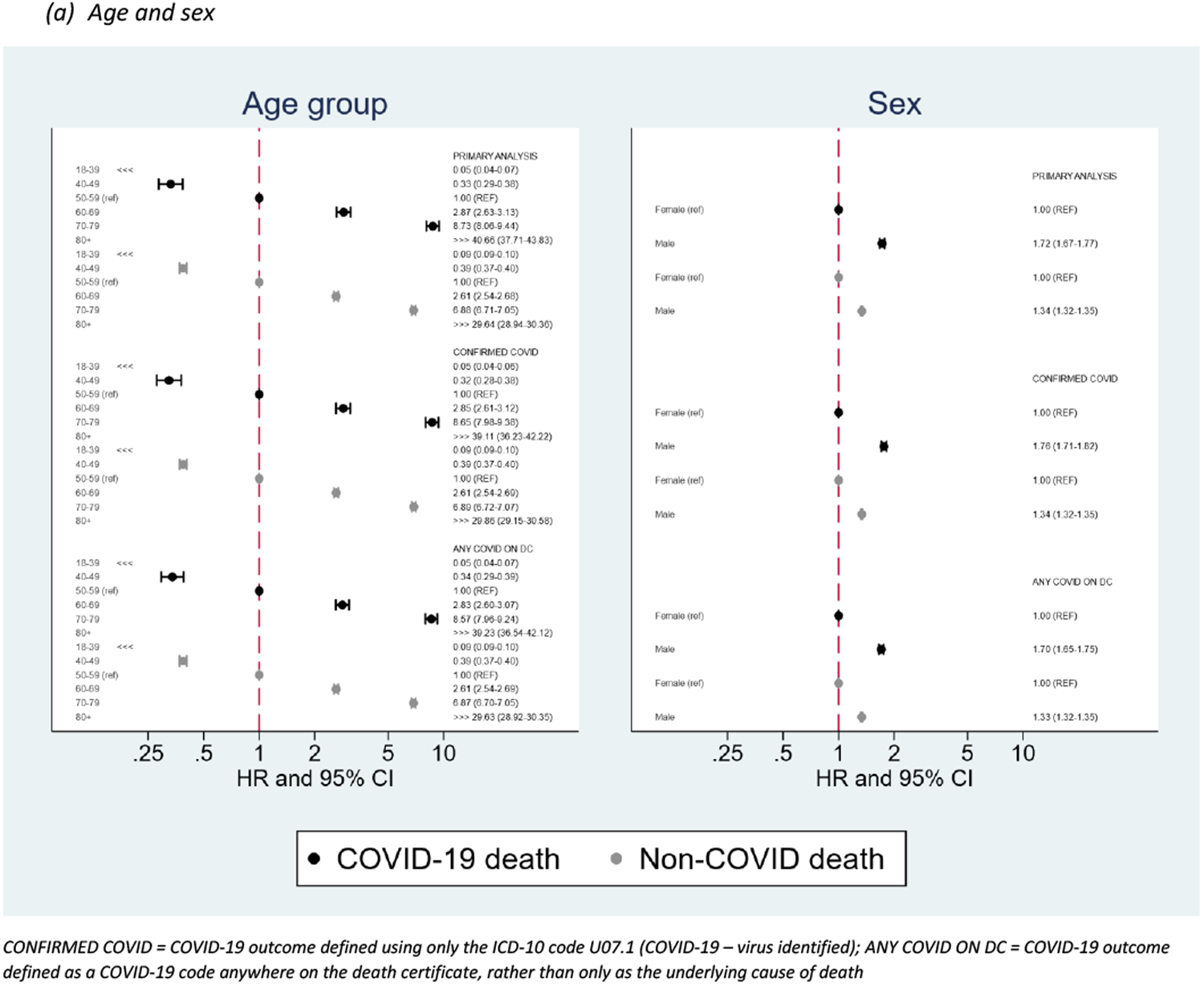
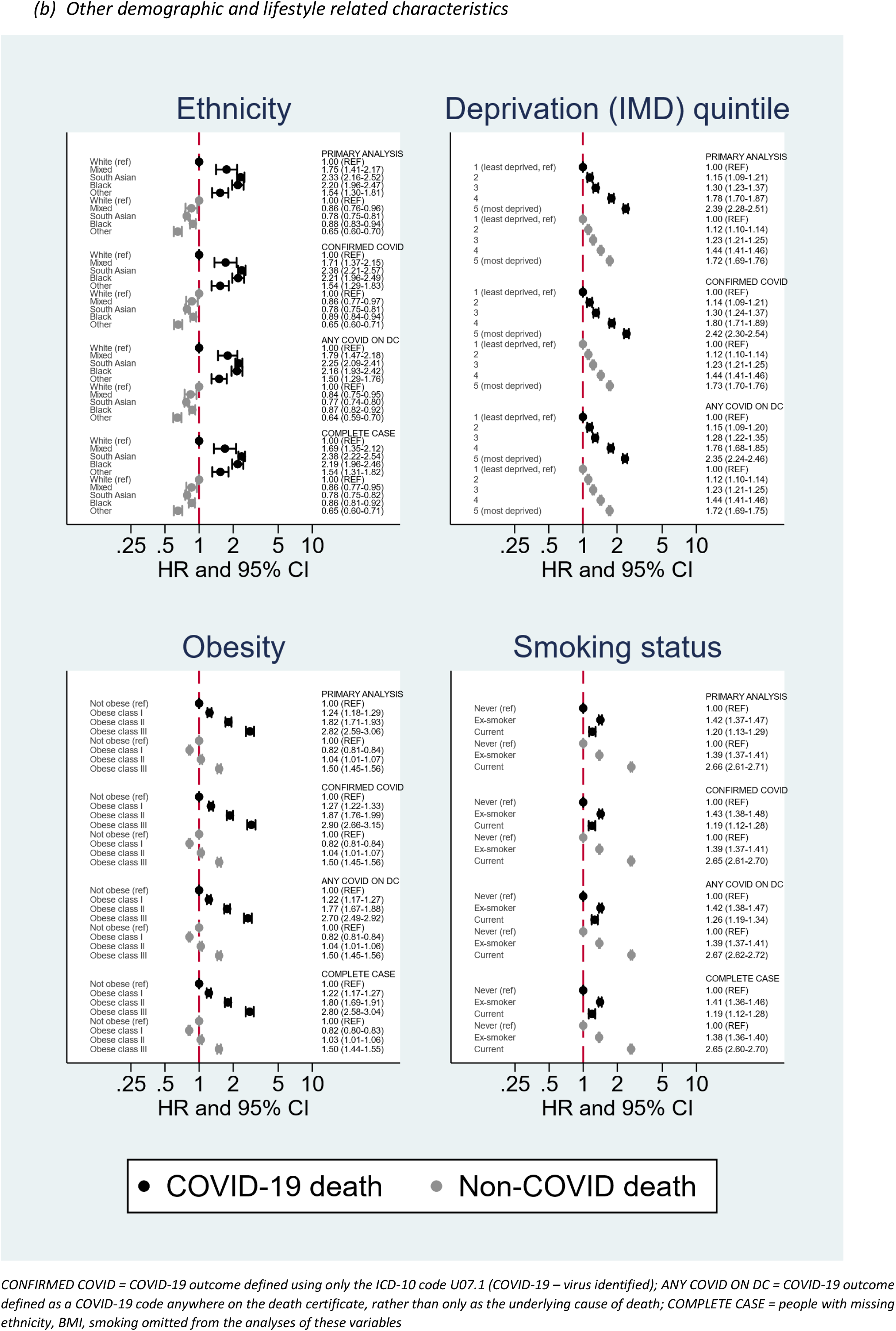
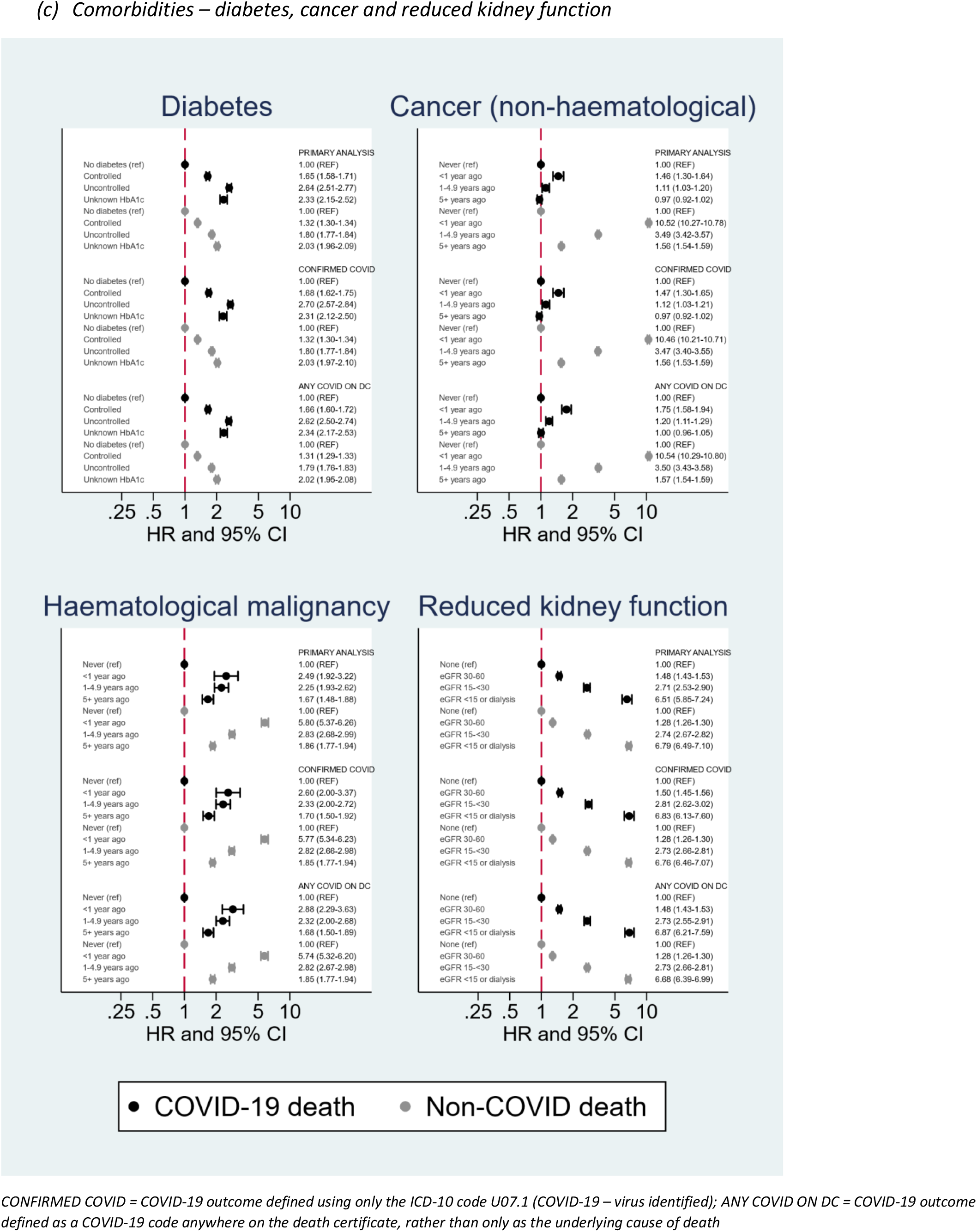
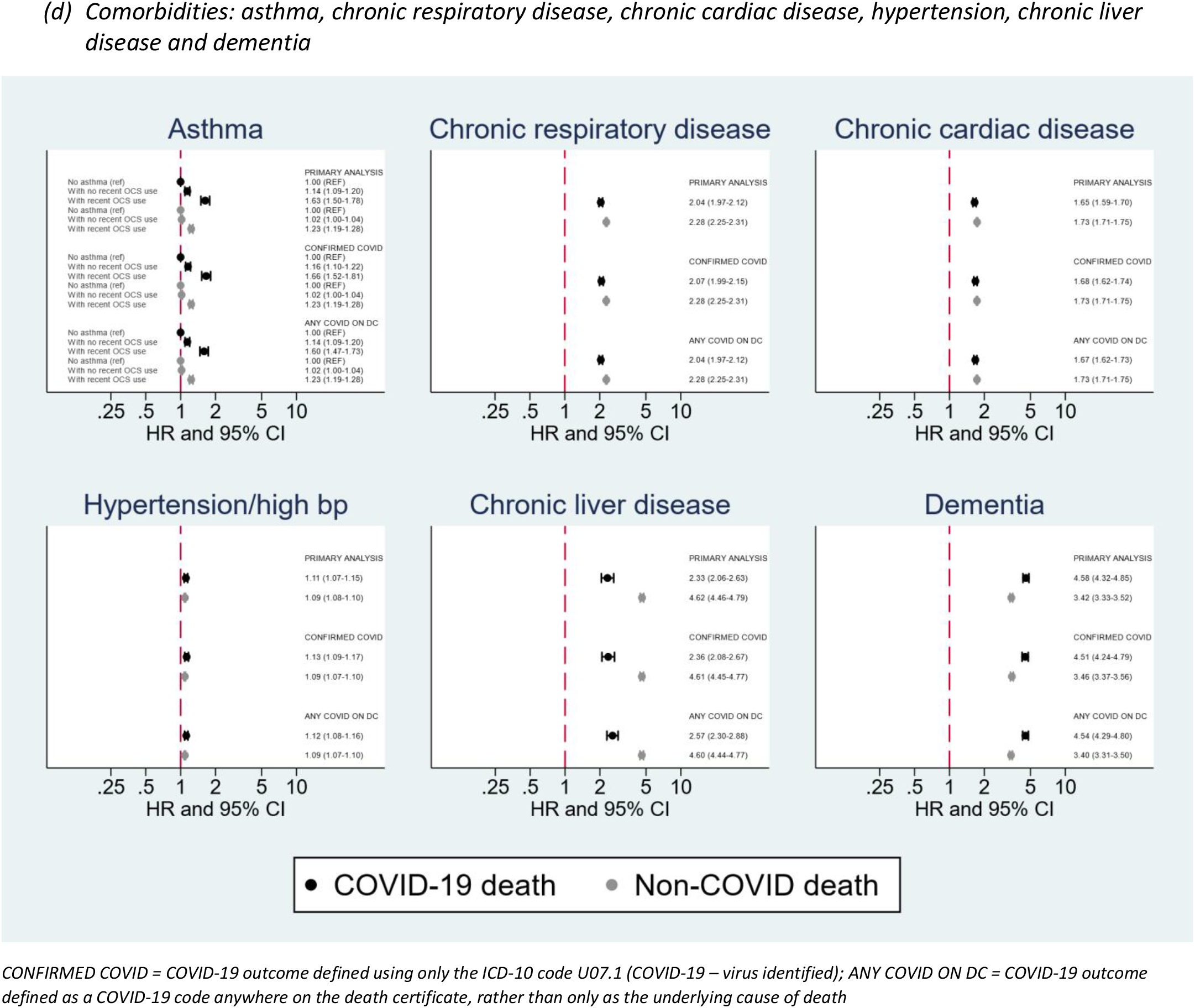
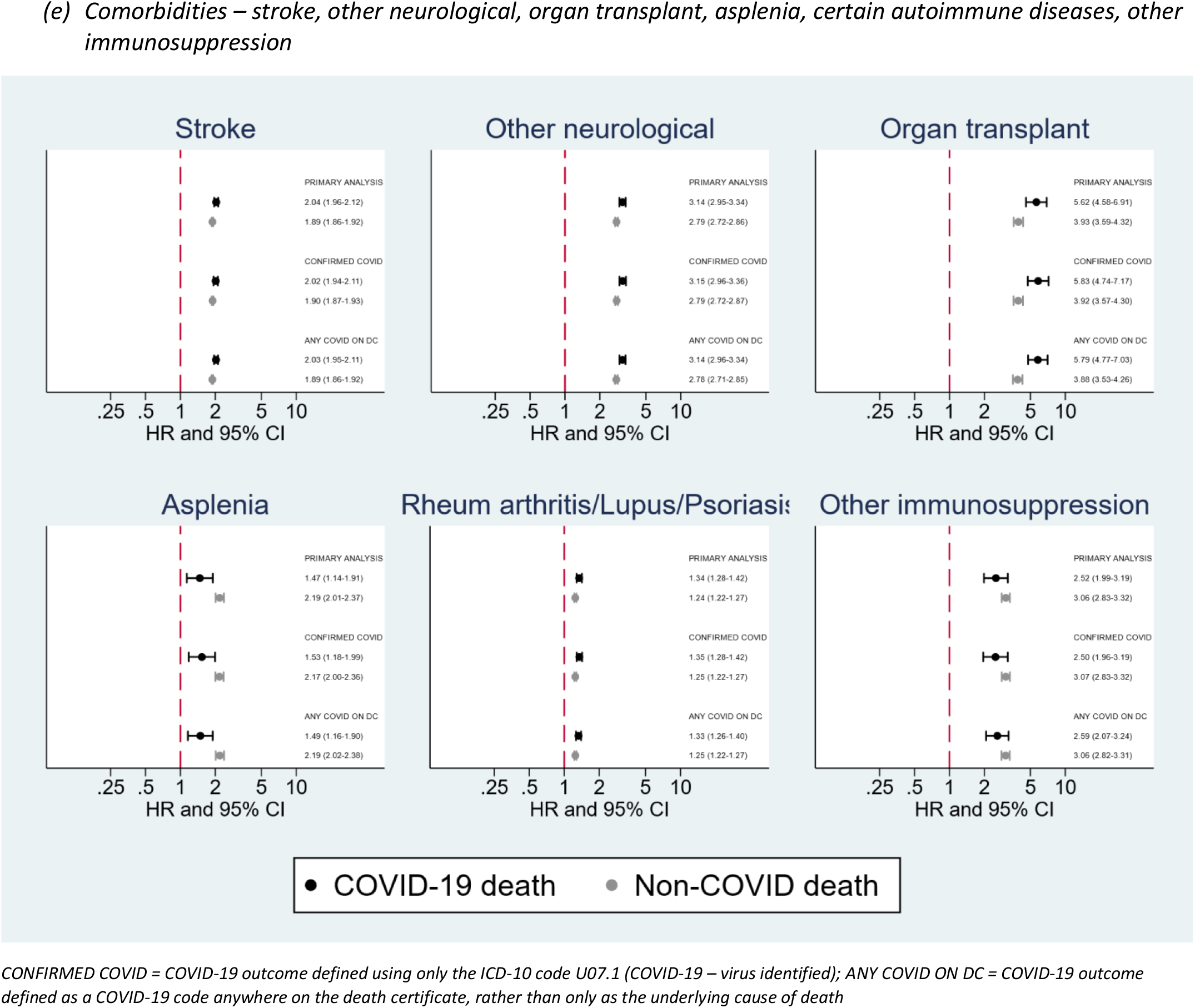
Age-sex adjusted odds ratios for associations between individual-level factors and COVID-19/non-COVID deaths in sensitivity analyses

## References

1. European Centre for Disease Prevention and Control. COVID-19 situation update worldwide, as of 26 November 2020. 2020. https://www.ecdc.europa.eu/en/geographical-distribution-2019-ncov-cases (accessed 26 November 2020).

2. Zhou F, Yu T, Du R, Fan G, Liu Y, Liu Z, Xiang J, Wang Y, Song B, Gu X. Clinical course and risk factors for mortality of adult inpatients with COVID-19 in Wuhan, China: a retrospective cohort study. The lancet 2020. doi.

3. Williamson EJ, Walker AJ, Bhaskaran K, Bacon S, Bates C, Morton CE, Curtis HJ, Mehrkar A, Evans D, Inglesby P. OpenSAFELY: factors associated with COVID-19 death in 17 million patients. Nature 2020: 1–11. doi: 10.1038/s41586-020-2521-4.

4. Mathur R, Rentsch CT, Morton C, Hulme WJ, Schultze A, MacKenna B, Eggo RM, Bhaskaran K, Wong AY, Williamson EJ, Forbes H, Wing K, McDonald HI, Bates C, Bacon S, Walker AJ, Evans D, Inglesby P, Mehrkar A, Curtis HJ, DeVito NJ, Croker R, Drysdale H, Cockburn J, Parry J, Hester F, Harper S, Douglas IJ, Tomlinson L, Evans S, Grieve R, Harrison D, Rowan K, Khunti K, Chaturvedi N, Smeeth L, Goldacre B. Ethnic differences in COVID-19 infection, hospitalisation, and mortality: an OpenSAFELY analysis of 17 million adults in England. medRxiv 2020: 2020.09.22.20198754. doi: 10.1101/2020.09.22.20198754.

5. Public Health England. Disparities in the risk and outcomes of COVID-19. 2020. doi.

6. Rentsch CT, Kidwai-Khan F, Tate JP, Park LS, King Jr JT, Skanderson M, Hauser RG, Schultze A, Jarvis CI, Holodniy M. Patterns of COVID-19 testing and mortality by race and ethnicity among United States veterans: A nationwide cohort study. PLoS medicine 2020; 17(9): e1003379. doi.

7. Spiegelhalter D. Use of “normal” risk to improve understanding of dangers of covid-19. BMJ 2020; 370: m3259. doi: 10.1136/bmj.m3259.

8. Drozd M, Lillie P, Pujades-Rodriguez M, Straw S, Morgan AW, Kearney MT, Witte KK, Cubbon RM. Non-communicable disease and the risk of infection death: a UK Biobank prospective cohort study. medRxiv 2020: 2020.07.27.20162354. doi: 10.1101/2020.07.27.20162354.

9. World Health Organisation. Emergency use ICD codes for COVID-19 disease outbreak. 2020. https://www.who.int/classifications/icd/covid19/en/ (accessed 6th August 2020).

10. Bhaskaran K, Smeeth L. What is the difference between missing completely at random and missing at random? International journal of epidemiology 2014; 43(4): 1336–9. doi: 10.1093/ije/dyu080.

11. NHS Digital. BETA – Data Security Standards. 2020. https://digital.nhs.uk/about-nhs-digital/our-work/nhs-digital-data-and-technology-standards/framework/beta---data-security-standards (accessed 6th August 2020).

12. NHS Digital. Data Security and Protection Toolkit. 2020. https://digital.nhs.uk/data-and-information/looking-after-information/data-security-and-information-governance/data-security-and-protection-toolkit (accessed 6th August 2020).

13. NHS Digital. ISB1523: Anonymisation Standard for Publishing Health and Social Care Data. 2020. https://digital.nhs.uk/data-and-information/information-standards/information-standards-and-data-collections-including-extractions/publications-and-notifications/standards-and-collections/isb1523-anonymisation-standard-for-publishing-health-and-social-care-data (accessed 6th August 2020).

14. Secretary of State for Health - UK Government. Coronavirus (COVID-19): notification to organisations to share information. 2020. https://www.gov.uk/government/publications/coronavirus-covid-19-notification-of-data-controllers-to-share-information (accessed 6th August 2020).

15. Bhopal RS, Gruer L, Cezard G, Douglas A, Steiner MF, Millard A, Buchanan D, Katikireddi SV, Sheikh A. Mortality, ethnicity, and country of birth on a national scale, 2001–2013: A retrospective cohort (Scottish Health and Ethnicity Linkage Study). PLoS medicine 2018; 15(3): e1002515. doi.

16. Fry A, Littlejohns TJ, Sudlow C, Doherty N, Adamska L, Sprosen T, Collins R, Allen NE. Comparison of Sociodemographic and Health-Related Characteristics of UK Biobank Participants With Those of the General Population. American Journal of Epidemiology 2017; 186(9): 1026–34. doi: 10.1093/aje/kwx246.

17. Office for National Statistics. Coronavirus (COVID-19) related deaths by ethnic group, England and Wales: 2 March 2020 to 15 May 2020. 2020. https://www.ons.gov.uk/peoplepopulationandcommunity/birthsdeathsandmarriages/deaths/articles/coronaviruscovid19relateddeathsbyethnicgroupenglandandwales/2march2020to15may2020 (accessed November 27 2020).

18. Simons D, Shahab L, Brown J, Perski O. The association of smoking status with SARS-CoV-2 infection, hospitalisation and mortality from COVID-19: A living rapid evidence review (version 5). Qeios 2020. doi: 10.32388/UJR2AW.6.

19. Carreira H, Strongman H, Peppa M, McDonald HI, dos-Santos-Silva I, Stanway S, Smeeth L, Bhaskaran K. Prevalence of COVID-19-related risk factors and risk of severe influenza outcomes in cancer survivors: A matched cohort study using linked English electronic health records data. EClinicalMedicine 2020; 29. doi: 10.1016/j.eclinm.2020.100656.

20. Kraindler J, Barclay C, Tallack C,. Understanding changes to all mortality during the pandemic. https://www.health.org.uk/news-and-comment/charts-and-infographics/understanding-changes-to-all-mortality-during-the-pandemic (accessed 22nd December 2020).

21. Fine JP, Gray RJ. A proportional hazards model for the subdistribution of a competing risk. J Am Stat Assoc 1999; 94(446): 496–509. doi: Doi 10.2307/2670170.

22. Clift AK, Coupland CAC, Keogh RH, Diaz-Ordaz K, Williamson E, Harrison EM, Hayward A, Hemingway H, Horby P, Mehta N, Benger J, Khunti K, Spiegelhalter D, Sheikh A, Valabhji J, Lyons RA, Robson J, Semple MG, Kee F, Johnson P, Jebb S, Williams T, Hippisley-Cox J. Living risk prediction algorithm (QCOVID) for risk of hospital admission and mortality from coronavirus 19 in adults: national derivation and validation cohort study. BMJ 2020; 371: m3731. doi: 10.1136/bmj.m3731.

23. Coggon D, Croft P, Cullinan P, Williams A. Assessment of workers’ personal vulnerability to covid-19 using ‘covid-age’. Occupational Medicine 2020; 70(7): 461–4. doi: 10.1093/occmed/kqaa150.

